# THE USE OF DENATURING SOLUTION AS COLLECTION AND TRANSPORT MEDIA TO IMPROVE SARS-COV-2 RNA DETECTION AND REDUCE INFECTION OF LABORATORY PERSONNEL

**DOI:** 10.1101/2020.06.18.20134304

**Authors:** Alex Fiorini de Carvalho, Andreza Parreiras Gonçalves, Thaís Bárbara de Souza Silva, Hugo Itaru Sato, Larissa Vuitika, Flavia Fonseca Bagno, Sarah Aparecida Rodrigues Sérgio, Maria Marta Figueiredo, Raissa Prado Rocha, Ana Paula Salles Moura Fernandes, Pedro Augusto Alves, Santuza Maria Ribeiro Teixeira, Flavio Guimarães da Fonseca

## Abstract

**Background:** Since the emergence of the COVID-19, health officials have struggled to devise strategies to counteract the speed of the pandemic’s spread across the globe. It became imperative to implement accurate diagnostic tests for the detection of SARS-CoV-2 RNA on respiratory samples. In many places, however, besides the limited availability of test reagents, laboratory personnel face the challenge of adapting their working routines to manipulate highly infective clinical samples. Here, we proposed the use of a virus-inactivating solution as part of a sample collection kit to decrease the infectious potential of the collected material without affecting the integrity of RNA samples used in diagnostic tests based on RT-qPCR.

**Methods:** Nasopharyngeal and oropharyngeal swab samples were collected from SARS-CoV-2-infected patients and from laboratory personnel using a commercially available viral transport solution (VTM) and the denaturing solution (DS) described here. RNA extracted from all samples was tested by RT-qPCR using probes for viral and human genes. Exposure of laboratory personnel to infective viruses was also accessed using ELISA tests.

**Findings:** The use of the DS did not interfere with the detection of viral genome or the endogenous human mRNA, since similar results were obtained from samples collected with VTM or DS. In addition, all tests of laboratory personnel for the presence of viral RNA and IgG antibodies against SARS-CoV-2 were negative.

**Interpretation:** The methodology described here provides a strategy that allow high diagnostic accuracy as well as safe manipulation of clinical samples by those involved with diagnostic procedures.

**Funding:** CAPES, FAPEMIG, CNPq, MCTIC, FIOCRUZ and the UK Global Challenges Research Fund (GCRF).

## 1. Introduction

In December 2019, Chinese health officials reported several cases of respiratory syndrome followed by pneumonia of unknown origin, initially in the city of Wuhan, capital of the Hubei province. The etiological agent behind the upsurge of the new syndrome was quickly identified as a new coronavirus, latter on named SARS-CoV-2.^1^ The virus spread rapidly throughout China and in less than a month reached other countries in Asia, eventually reaching other continents. On March 11, 2020, a global pandemic was declared by the World Health Organization (WHO).^2, 3^ To date, more than 6 million cases and more than 440 thousand deaths due to the disease caused by SARS-CoV-2, named COVID-19, have been recorded worldwide, in 188 countries and territories around the planet with thousands of new cases and deaths been reported every day.^4^

While health officials and governments around the world struggle to devise strategies to counteract the pace of the infection’s spread, efforts to implement fast and sensitive approaches for diagnostic have emerged as key steps to control the epidemics. Real-time reverse transcriptase-PCR (RT-PCR) testing to detect SARS-CoV-2 RNA on samples collected from the largest possible fraction of the populations became an absolute consensus.^5^ Widespread PCR testing has been pointed out as one of the most important elements in the successful COVID-19 containment strategy adopted by countries that have shown positive outcomes, including Taiwan, South Korea and Germany.^6, 7, 8^ Nonetheless, having test kits available is not the only bottleneck to implement universal testing in many countries. The capability to adapt the routines of diagnostic laboratories to cope with the manipulation of highly infective clinical samples coming by the thousands is equally essential, especially considering that the RT-PCR diagnostic requires highly trained laboratory personnel.

After peaking in Asiatic and European countries, the spread of the disease veered towards the Americas and possibly the sub-Saharan African continent. Developing countries in these continents may be hit hard by the pandemic for a number of reasons; and one particular troublesome aspect is the limited availability of diagnostic laboratories that will be able to cope with the huge incoming of clinical samples – either in terms of the total number of available laboratories, or their capability to deal safely with potentially infective clinical specimens. Therefore, the development of strategies to reduce the infectivity of clinical samples being sent to diagnostic laboratories could be essential to avoid contamination of the limited number of trained personnel and to maintain the operational capability of these laboratories.

The high risk associated with biological samples determines that any clinical samples are to be considered as potentially infectious and, therefore, must be treated under strict biosafety protocols.^9^ In this regard, national and international guidelines on biosafety concerning clinical laboratories must be followed in all circumstances. In the context of the current COVID-19 pandemic, there is still limited information regarding nosocomial infection by SARS-CoV-2 affecting health workers involved in diagnostics or similar activities. The WHO recommends that handling of clinical samples suspected of being infected with SARS-CoV-2 requires a BSL-2 or equivalent facility, whereas attempts to replicate the virus require at least BSL-3 facilities.^10^

Several chemical and physical methods of viral inactivation have been proposed and evaluated for different pathogens, as a way to provide greater safety for professionals involved in the handling of potentially infectious samples and lower costs with laboratory infrastructure.^9, 11, 12, 13, 14, 15, 16^ Amongst the chemical methods evaluated, the most commonly used products contain a chaotropic salt (guanidine), which acts as a denaturant agent for macromolecules culminating in virus inactivation.^12^ At the same time, guanidine is able to decrease the degradation of RNA molecules in samples, acting as a ribonuclease inhibitor, therefore increasing the preservation of genetic material for application in molecular diagnostic methodologies in which RNA integrity is essential.^13^

Here we describe the use of a simple, virus-inactivating and denaturing solution as part of a swab collection kit, aiming to decrease the infectious potential of the clinical sample and, at the same time, to preserve highly frail RNA molecules during transportation and short-term storage before testing. This low-cost, accessible approach has made it possible to achieve high diagnostic accuracy as well as manipulation safety for those involved with diagnostic procedures.

## 2. Methodology

### 2.1. Denaturing Solution (DS)

The guanidine isothiocyanate solution used in this study is based on protocols established by Zolfaghari et al.,^17^ and Chomczynski and Sacchi^18^. The denaturing solution was prepared using 4M guanidine isothiocyanate, 2.5M sodium citrate and RNAse/DNAse free water and pH adjusted to 4.0. This solution has been used for the conditioning and transportation of clinical specimens that include oropharyngeal and nasopharyngeal swabs. The guanidine chaotropic salt was chosen as the denaturing agent, assuming its ability to inactivate viruses.^12^ In addition, guanidine is also a ribonuclease inhibitor, which allows the conservation of genetic material for downstream applications using molecular biology methodologies.^19^

### 2.2. Molecular diagnostic

#### 2.2.1. Sampling

When the SARS-CoV2 was declared pandemic, in early march 2020, research groups from major Universities and research institutes throughout Brazil begin to prepare themselves to offer diagnostic support, correctly foreseeing the collapse of public and private laboratories’ capabilities shortly after the epidemics reached the Country. At the UFMG’s Vaccine Technology Center, we were particularly concerned about the impacts of the intense flow of infective samples in a research laboratory that was adapted to join the testing effort with limited resources and personnel. In order to increase personnel safety, to avoid losing collaborators due to infections by SARS-CoV-2, and at the same time to increase preservation of the RNA contained in clinical samples, we introduced the use of the guanidine-containing solution as collection and transport media instead of commonly used viral transport media (VTM). VTM is usually composed of a balanced salt buffer; sterile, heat-inactivated fetal bovine serum; and antibiotics, as suggested by the CDC.^20^ As recommended in the WHO interim guidance protocol^10^, combined oropharyngeal and nasopharyngeal swabs were collected, using sterile flexible-rod swabs, and placed in a single sterile 15 mL polypropylene tube, containing 1.0 mL of the described denaturing solution. These collection kits were prepared in our laboratory and sent to hospitals according to their daily demand. Upon sample collection, the swabs remained immersed in the denaturing solution for at least 30 seconds, after which they were removed while being gently pressed against the tube wall to remove the excess absorbed solution. Swabs were, discarded in an appropriated biological waste disposal and were not sent to the diagnostic laboratory in order to minimize risks of personnel contamination. Clinical specimens from the laboratory personnel were collected multiple times and processed the same way as specimens from patients in hospitals (see below).

#### 2.2.2. RNA extraction and Real-Time Polymerase Chain Reaction (qPCR)

Extraction of the total RNA from samples was performed using the QIAamp Viral RNA Mini Kit (Qiagen, USA), according to protocols provided by the manufacturer. The real time polymerase chain reaction (qPCR) was performed using primers and probes described in two different protocols: the United States Center for Disease Control and Prevention (CDC) protocol^20^ and the Berlin (Charité/Berlin) protocol.^21^ The viral gene coding for the N protein was targeted for the detection of SARS-CoV-2 RNA using the CDC protocol, and the viral gene E was used in the Charité/Berlin protocol. Probes and primers for the human RNAse P mRNA were used in both protocols as an endogenous reaction control. Reactions were carried out with the Promega GoTaq® Probe 1-Step RT-qPCR Kit (Promega, France) according to the manufacturer’s recommendations, using the QuantStudio™ 5 Real-Time PCR System (Thermo Fisher, USA).

### 2.3. RNA Viability Test

In order to verify the viability of the RNA stored in the denaturing solution (DS) proposed in this work, in comparison to the commonly used VTM, oropharyngeal and nasopharyngeal swab samples were collected from four laboratory’s staff members and stored in VTM, following the protocol established by the WHO. Similarly, swab samples from the same staff were placed in the DS. After a two hours period, RNA extraction was performed from all samples, as described above, and the qPCR protocol was performed to detect the RNAse P gene. To evaluate for how long the DS could maintain viral RNA viable to be detected through qPCR, different DS tubes were spiked with 16400 copies of SRAS-CoV2 genomic RNA and maintained for up to 8 days at either 4°C or room temperature. After that, the viral RNA was extracted and evaluated by qPCR. Results obtained were analyzed in the QuantStudio™ Design and Analysis software (v.1.5.1) and graphs were generated using the GraphPad Prism software (v.8.4.2).

### 2.4. Detection of Viral RNA from samples stored in the Viral Transport Medium and the Denaturing Solution

To evaluate the preservation of the viral RNA in VTM versus DS, clinical samples were collected from hospitalized COVID-19 suspected patients using either DS or VTM. Sample collection was carried out according to the WHO protocol and the molecular diagnosis was processed as described above. Obtained results were analyzed in the QuantStudio™ Design and Analysis software (v.1.5.1) and graphs were generated using the GraphPad Prism software (v.8.4.2).

### 2.5. Assessment of laboratory personnel safety

Having established that collection of clinical samples in DS preserves viral RNA in levels comparable to VTM, we opted to routinely receive and process only DS collected clinical samples, as described above. In order to assess the safety of our laboratory staff using such routine, we tested all laboratory personnel every 15 days. qPCR tests were conducted as described above. Additionally, serum samples from all laboratory members were also collected approximately 45 days after the beginning of the study, to assess the presence of antibodies against SARS-CoV-2. To that end, we employed an in-house anti-IgG COVID-19 enzyme-linked immunosorbent assay. Briefly, the nucleocapsid (N) protein of SARS-CoV-2 was expressed in transformed *E. coli*, purified, and used to coat 96 well ELISA plates. Tested sera were diluted in PBS-Tween20 solution, added to wells and incubated for 1 hour at 37°C. After washing, each well were added with horseradish peroxidase (HRP)-conjugated anti-human IgG goat immunoglobulin (Fapon, China). After further washing and incubation, reactions were revealed using tetramethylbenzidine (TMB) and readings were obtained in a Microplate Reader at an optical density (O.D.) of 450 nm. The in-house indirect ELISA was validated using a panel of SARS-CoV-2-positive sera previously tested by commercial anti-SARS-CoV-2 IgG lateral flow immunochromatography (various commercial brands).

### 2.6. Ethical Considerations

This work was approved by the UFMG’s Ethics Committee and by the National Research Ethics’ Committee, under number CAAE 31686320.0.000.5149. All laboratory personnel signed informed consents.

## 3. Results

### 3.1. RNA Viability Tests

Oropharyngeal and nasopharyngeal samples from four different laboratory members were collected in VTM or DS and processed. Viability of the extracted RNA was analyzed by qPCR, looking for the detection of the RNAse P human mRNA. We observed no differences in the cycle threshold (C_T_) obtained for the detection of the targeted mRNA, regardless of the collection media (samples A to D), suggesting that the potential preservation of RNA in the two solutions is similar (Figure 1).

**Figure 1.**
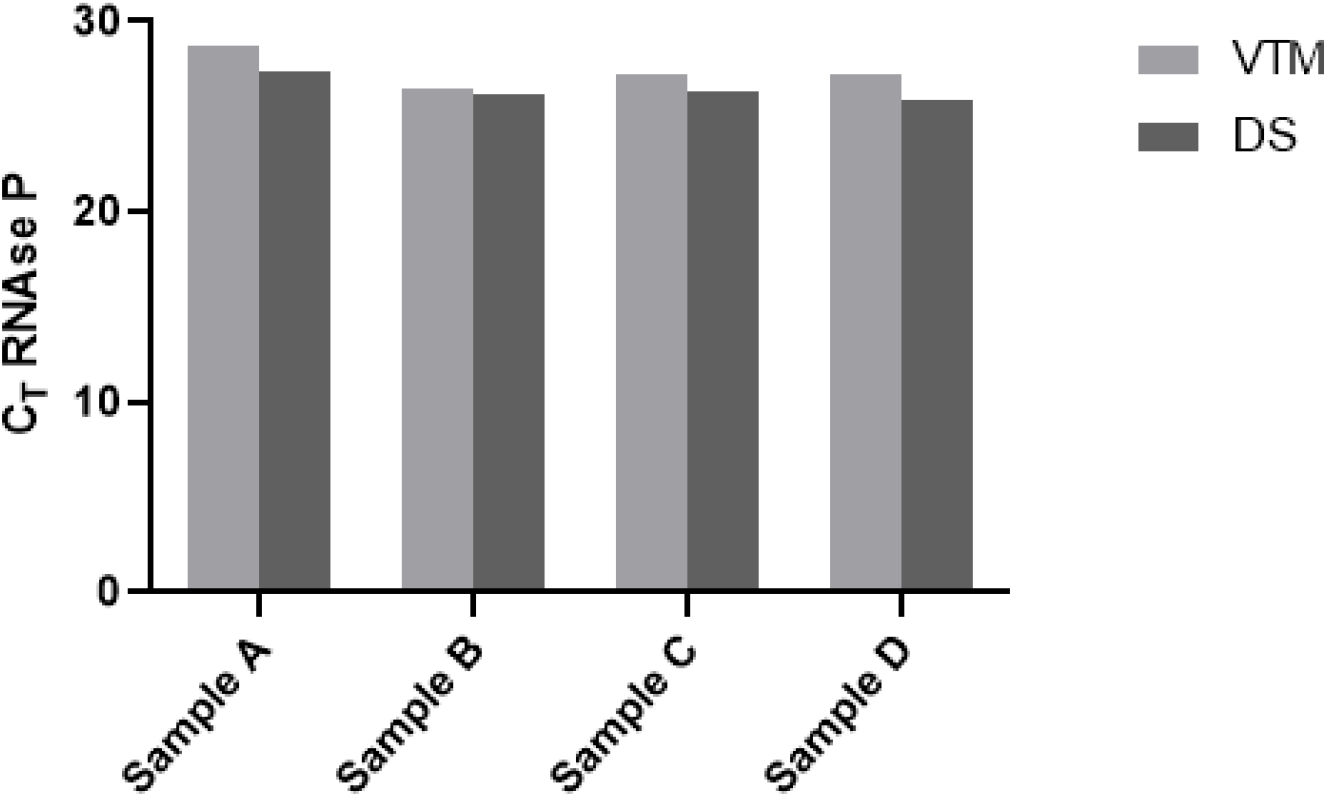
Comparative analysis of RNAse P mRNA amplification in samples extracted from VTM and DS. C_T_ obtained for the RNAse P mRNA from samples A, B, C and D, stored and extracted from VTM (virus transport media) or DS (denaturing solution).

Having established that DS can be reliably used to collect genetic material, we next asked for how long DS would keep viral genomic RNA viable for detection, either at 4°C or at room temperature. When compared to day 0 after spiking SARS-Cov2 RNA into DS tubes, there were no differences in viral RNA detection in days 1, 2, 4, 8 and 16 stored under refrigeration or at room temperature (Figure 2).

**Figure 2.**
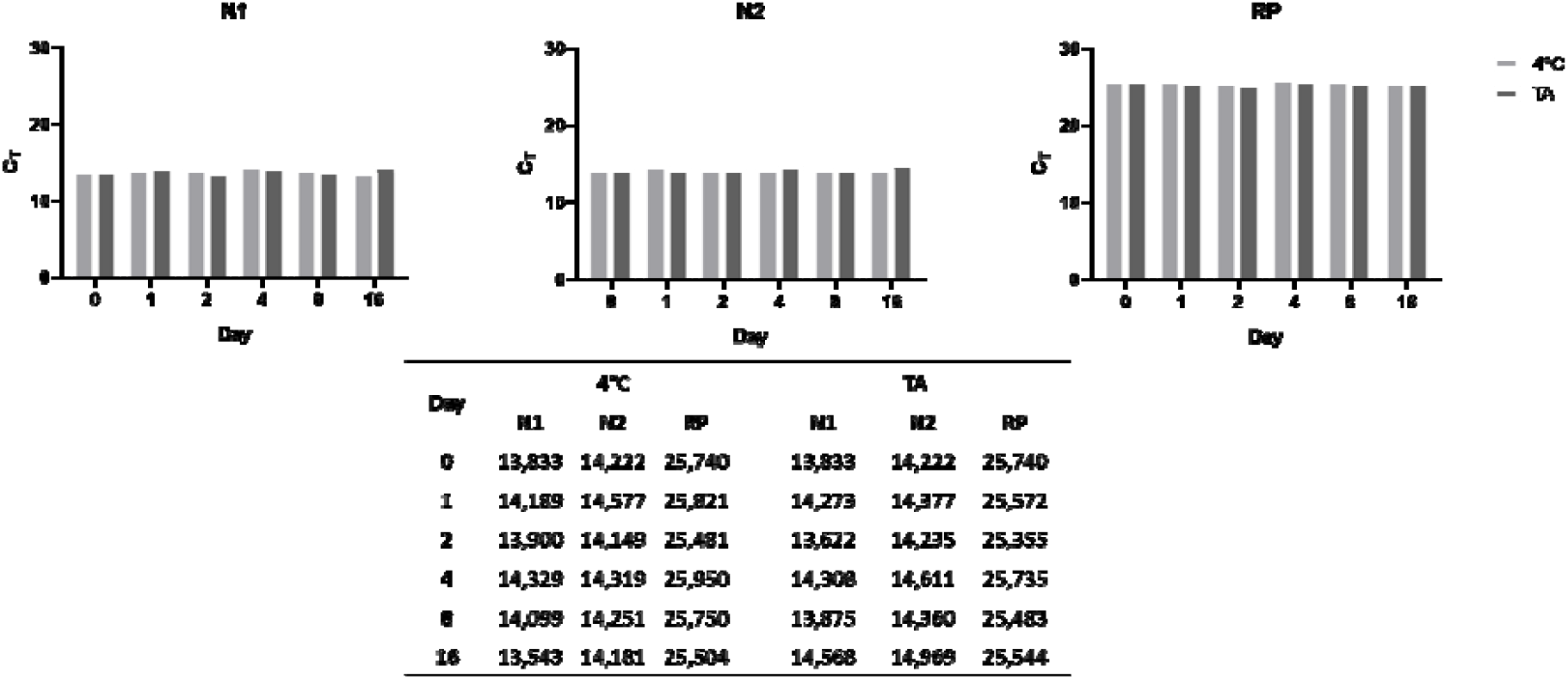
RNA Viability in DS solution after storage at 4°C and room temperature for 16 days. DS tubes were spiked with SARS-Cov2 RNA and stored under refrigeration or at room temperature. The presence of SARS-Cov2 RNA was detected by qPCR (N1 and N2 viral genes) in the samples after 1, 2, 4, 8, and 16 days of storage.

### 3.2. Detection of Viral RNA from samples stored in the Viral Transport Medium and the Denaturing Solution

We next evaluated the detection of SARS-CoV-2 viral RNA from clinical samples collected in VTM or DS. We observed no difference in the amplification profile (determined by the cycle threshold) of the N1, N2, N3 viral genes or of the RNAse P mRNA (endogenous control), regardless of the employed collection media (Figure 3). Results depicted on figure 3 show the mean C_T_ from six randomly selected SARS-CoV-2-positive patients compared to six randomly selected SARS-CoV-2-negative individuals. These results are representative of a much larger panel of results obtained so far. All tests were also conducted using the Charité/Berlin qPCR protocol and results were similar (not shown).

**Figure 3.**
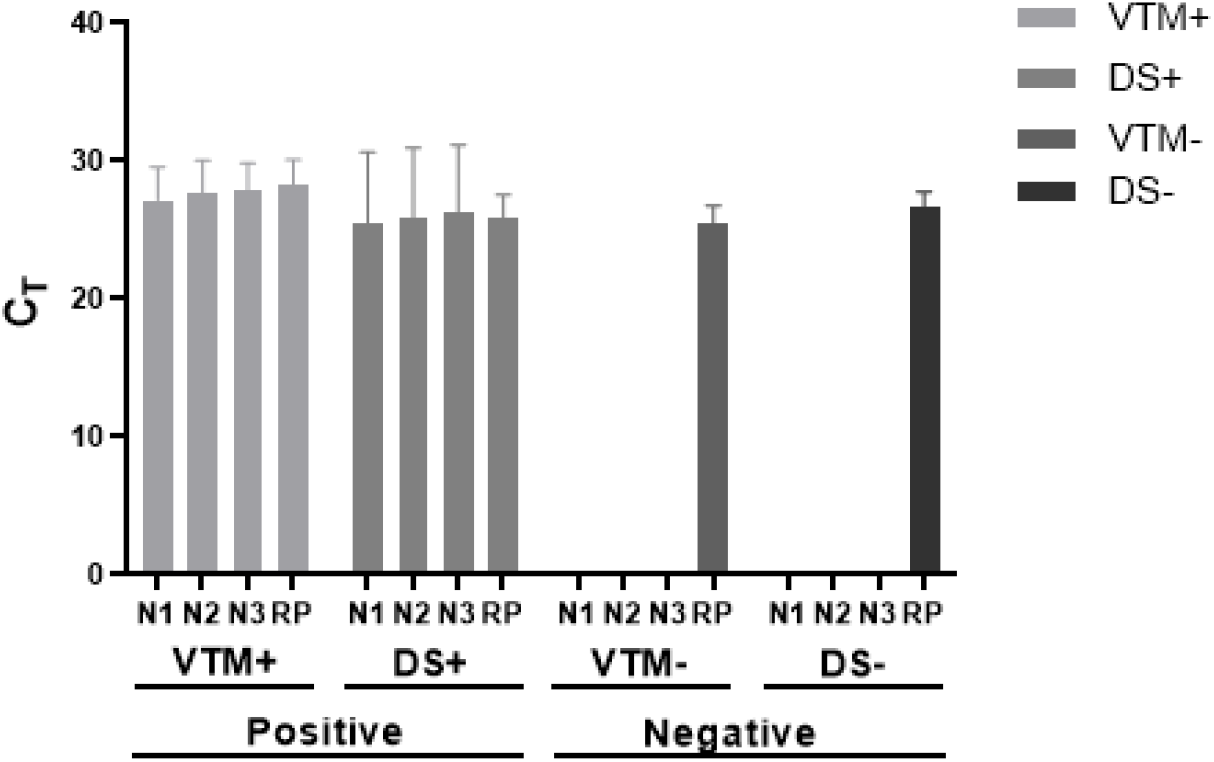
Comparative analysis of viral genes and RNAse P mRNA amplification in clinical samples extracted from VTM or DS. Mean C_T_ of positive and negative samples for N1, N2 and N3 viral genes, extracted from VTM and DS (n=6). VTM+: positives samples with viral transport media, DS+: positives samples with denaturing solution, VTM-: negative samples with viral transport media, DS-: negative samples with denaturing solution.

### 3.3. Assessment of laboratory personnel safety

All 19 laboratory personnel working at the UFMG’s Vaccine Technology Center were evaluated throughout the COVID-19 epidemic in Brazil, since March 2020, when our laboratory started to offer diagnostic support to hospitals and other public and private diagnostic laboratories. Laboratory members were tested at least 4 times (with one exception), in 15 days interval, and also by an in-house anti-IgG COVID-19 ELISA. Notably, none of the laboratory members tested positive in any of the tests (Table 1). PCR tests were also conducted using the Charité/Berlin protocol and results were similar (not shown). These findings indicate that there was no SARS-CoV-2 contamination of any of the professionals involved in the diagnostic process.

**Table 1.**
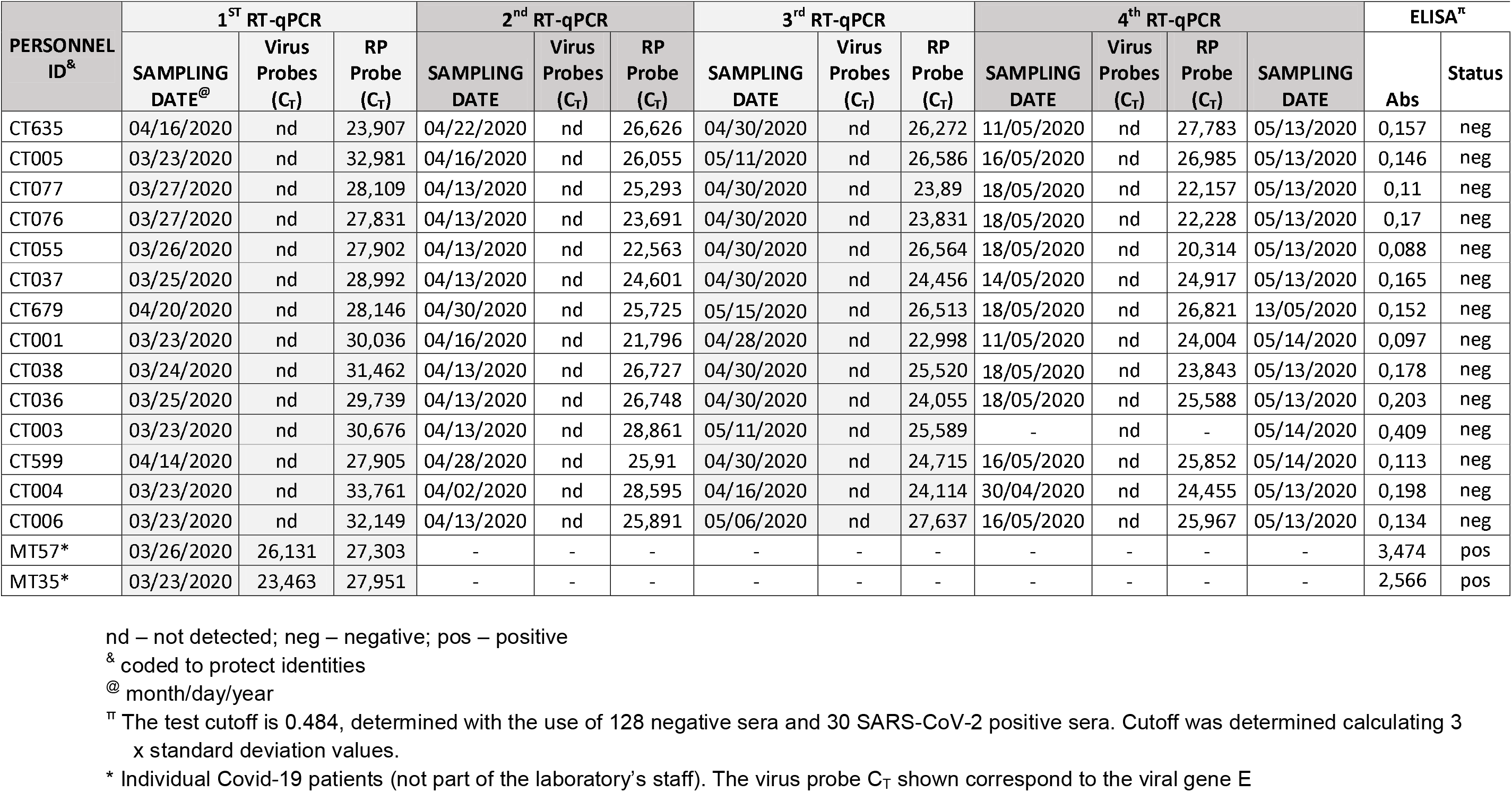

| PERSONNEL ID<sup>&</sup> | 1<sup>st</sup> RT-qPCR |  |  | 2<sup>nd</sup> RT-qPCR |  |  | 3<sup>rd</sup> RT-qPCR |  |  | 4<sup>th</sup> RT-qPCR |  |  |  | ELISA<sup>π</sup> |  |
| --- | --- | --- | --- | --- | --- | --- | --- | --- | --- | --- | --- | --- | --- | --- | --- |
|  | SAMPLING DATE<sup>@</sup> | Virus Probes (C<sub>T</sub>) | RP Probe (C<sub>T</sub>) | SAMPLING DATE | Virus Probes (C<sub>T</sub>) | RP Probe (C<sub>T</sub>) | SAMPLING DATE | Virus Probes (C<sub>T</sub>) | RP Probe (C<sub>T</sub>) | SAMPLING DATE | Virus Probes (C<sub>T</sub>) | RP Probe (C<sub>T</sub>) | SAMPLING DATE | Abs | Status |
| CT635 | 04/16/2020 | nd | 23,907 | 04/22/2020 | nd | 26,626 | 04/30/2020 | nd | 26,272 | 11/05/2020 | nd | 27,783 | 05/13/2020 | 0,157 | neg |
| CT005 | 03/23/2020 | nd | 32,981 | 04/16/2020 | nd | 26,055 | 05/11/2020 | nd | 26,586 | 16/05/2020 | nd | 26,985 | 05/13/2020 | 0,146 | neg |
| CT077 | 03/27/2020 | nd | 28,109 | 04/13/2020 | nd | 25,293 | 04/30/2020 | nd | 23,89 | 18/05/2020 | nd | 22,157 | 05/13/2020 | 0,11 | neg |
| CT076 | 03/27/2020 | nd | 27,831 | 04/13/2020 | nd | 23,691 | 04/30/2020 | nd | 23,831 | 18/05/2020 | nd | 22,228 | 05/13/2020 | 0,17 | neg |
| CT055 | 03/26/2020 | nd | 27,902 | 04/13/2020 | nd | 22,563 | 04/30/2020 | nd | 26,564 | 18/05/2020 | nd | 20,314 | 05/13/2020 | 0,088 | neg |
| CT037 | 03/25/2020 | nd | 28,992 | 04/13/2020 | nd | 24,601 | 04/30/2020 | nd | 24,456 | 14/05/2020 | nd | 24,917 | 05/13/2020 | 0,165 | neg |
| CT679 | 04/20/2020 | nd | 28,146 | 04/30/2020 | nd | 25,725 | 05/15/2020 | nd | 26,513 | 18/05/2020 | nd | 26,821 | 13/05/2020 | 0,152 | neg |
| CT001 | 03/23/2020 | nd | 30,036 | 04/16/2020 | nd | 21,796 | 04/28/2020 | nd | 22,998 | 11/05/2020 | nd | 24,004 | 05/14/2020 | 0,097 | neg |
| CT038 | 03/24/2020 | nd | 31,462 | 04/13/2020 | nd | 26,727 | 04/30/2020 | nd | 25,520 | 18/05/2020 | nd | 23,843 | 05/13/2020 | 0,178 | neg |
| CT036 | 03/25/2020 | nd | 29,739 | 04/13/2020 | nd | 26,748 | 04/30/2020 | nd | 24,055 | 18/05/2020 | nd | 25,588 | 05/13/2020 | 0,203 | neg |
| CT003 | 03/23/2020 | nd | 30,676 | 04/13/2020 | nd | 28,861 | 05/11/2020 | nd | 25,589 | - | nd | - | 05/14/2020 | 0,409 | neg |
| CT599 | 04/14/2020 | nd | 27,905 | 04/28/2020 | nd | 25,91 | 04/30/2020 | nd | 24,715 | 16/05/2020 | nd | 25,852 | 05/14/2020 | 0,113 | neg |
| CT004 | 03/23/2020 | nd | 33,761 | 04/02/2020 | nd | 28,595 | 04/16/2020 | nd | 24,114 | 30/04/2020 | nd | 24,455 | 05/13/2020 | 0,198 | neg |
| CT006 | 03/23/2020 | nd | 32,149 | 04/13/2020 | nd | 25,891 | 05/06/2020 | nd | 27,637 | 16/05/2020 | nd | 25,967 | 05/13/2020 | 0,134 | neg |
| MT57* | 03/26/2020 | 26,131 | 27,303 | - | - | - | - | - | - | - | - | - | - | 3,474 | pos |
| MT35* | 03/23/2020 | 23,463 | 27,951 | - | - | - | - | - | - | - | - | - | - | 2,566 | pos |

nd – not detected; neg – negative; pos – positive
<sup>&</sup> coded to protect identities
<sup>@</sup> month/day/year
<sup>π</sup> The test cutoff is 0.484, determined with the use of 128 negative sera and 30 SARS-CoV-2 positive sera. Cutoff was determined calculating 3 x standard deviation values.
\* Individual Covid-19 patients (not part of the laboratory's staff). The virus probe C<sub>T</sub> shown correspond to the viral gene E

## 4. Discussion

Inactivation of potentially infectious clinical samples is a critically important process during all stages of sample manipulation, from the initial sample collection, to processing and the final discard of the residual material.^12^ Previous studies with other coronaviruses have established that treatment with heat, ultraviolet light, inactivating chemicals and a variety of detergents are effective in inactivating beta-coronavirus.^11, 22, 23, 24, 25^ Leclercq et al.,^26^ showed that heat treatment effectively inactivates MERS-CoV, and Kumar et al.^13^, demonstrated that the use of solutions containing guanidine isothiocyanate is capable of inactivating a strain of the MERS-CoV, closely related to SARS-CoV and SARS-CoV-2. At the same time, the use of denaturing agents such as guanidine isothiocyanate provides stability to the genetic material and has been largely used in protocols designed to purify nucleic acids.

In the present study, we have replaced the virus transport media (VTM) which is commonly used in diagnostic protocols ^27^ by a denaturing solution (DS) containing guanidine isothiocyanate, to improve conditions of transportation and storage of clinical samples suspected of being infected with SARS-CoV-2. Our data demonstrated that the use of the denaturing solution in the pre-analytical process does not interfere with the detection of the presence of viral genes or the endogenous human mRNA (RNAse P), and results obtaining from samples collected in VTM or DS were identical in terms of diagnostic accuracy. Of particular importance is the fact that the DS is able to keep RNA samples viable to be detected by qPCR for up to 16 days, either at 4°C or at room temperature. This is an essential feature considering the logistics to take collected samples to diagnostic laboratories in countries with limited numbers of qPCR-capacitated facilities. Also important, the multiple evaluation of all laboratory personnel demonstrated that there was no nosocomial infection or significant exposition to SARS-CoV-2 during more than two months working with an average of 80 samples/day, since the beginning of the diagnostic procedures conducted at the laboratory up to the present date. During this period, more than 3,000 tests were conducted, including samples from more than 2,100 patients and a SARS-CoV-2 positivity rate of approximately 25%.

The use of VTM is particularly indicated when virus viability is important, especially when SARS-CoV-2 isolation is to be attempted. However, only BSL-3 laboratories should be used to perform experiments involving replicative viruses, whereas diagnostic procedures that does not involve virus replication can be conducted in BSL-2 laboratories.^10^ Media containing live viruses undoubtedly brings risks to laboratory personnel that manipulate clinical samples under lower biosafety standards.^28^ Indeed, nosocomial exposition to SARS-CoV-2 in medical and laboratory personnel has been reported.^29, 30^ Therefore, the use of collection and transport media able to inactivate SARS-CoV-2 with no loss of the diagnostic analytical power can be critical to avoid nosocomial infections in such laboratories. This is particularly desirable, as the pandemic epicenter is moving from Asia, Europe and North America to South America and Africa, where diagnostic laboratories with adequate biosafety structures are much scarcer. In addition, the use of a denaturing solution to collect and transport clinical samples, as the one described here, reduces costs in the processing of samples.^11, 12, 13^

Our study have important limitations. First, this was not a case-control study, as our results were not compared to those obtained in a diagnostic laboratory routinely receiving clinical samples in VTM. Nonetheless, the differences in the possible extent of live virus exposition when VTM or DS are used are obvious. In this regard, the fact that none of our laboratory members was either infected or even seroconverted is an important indication that DS has been helpful in avoiding nosocomial exposition. Another limitation is that we are not able to quantify to which extent the good laboratory practices adopted in the laboratory could also be responsible for the verified results. Nonetheless, the use of the denaturing transport media is a critical part of such practices. Finally, we attempted to quantify the extent of SARS-CoV-2 inactivation using the described DS. To that end, isolated, laboratory cultivated live viruses were loaded on VTM or DS and plated on VERO cells. Viruses loaded on VTM remained replicative and able to generate plaques in the cellular monolayers (not shown). On the other hand, even when highly diluted, the guanidine salt present in DS was extremely toxic to cells, and the monolayers were destroyed before any eventual plaque could form. Nevertheless, it seems obvious to assume that, like cells, viruses were equally inactivated during exposition to DS. In spite of the limitations of our study, the use of denaturing, virus-inactivation solution as a collection and transportation media for diagnostic purposes is clearly an important asset to maximize clinical sample viability and minimize nosocomial infections in diagnostic laboratories, especially considering the SARS-CoV-2 spread to developing countries in which biosafe-structured laboratories are not easily available.

## Data Availability

All data referred to in the manuscript is fully available.

## 5 Acknowledgements

We thank all personnel at UFMG’s Vaccine Technology Center (CTVacinas) for their dedication to the COVID-19 diagnostic support in Belo Horizonte, Brazil. Members of the CTVacinas are composed of undergraduate and graduate students, post-doc fellows, professors and researchers at the Federal University of Minas Gerais and the René Rachou Institute (FIOCRUZ), who interrupted their research activities to dedicate themselves to aid people during such difficult times. We thank Anna Raquel dos Santos, Júlia Reis, Lisiane Gomes, Natalia Salazar and Ricardo Gazzinelli for their help and expertise. We thank Professor Édison L. Durigon, at Laboratório de Virologia Clínica e Molecular, ICB/USP, São Paulo, Brazil, who kindly provided inactivated samples of the virus isolate HIAE-02: SARS-CoV-2/SP02/human/2020/BRA (GenBank accession number MT126808.1). We also thank the Laboratório de Virus’ team, at ICB/UFMG, for valuable technical help. The research leading to these results has, in part, received funds from CAPES, CNPq, FAPEMIG, FIOCRUZ and from the MCTIC through the “*Rede Virus*” initiative. Funds came also from UK Research and Innovation via the Global Challenges Research Fund under grant agreement ‘A Global Network for Neglected Tropical Diseases’ (grant number MR/P027989/1). Further funding included Institutional financial resources of the René Rachou Institute - Oswaldo Cruz Foundation (FIOCRUZ) and the Instituto Nacional de Ciência e Tecnologia de Vacinas (INCTV).

